# Development and Content Validation of the Symptoms Evolution of COVID-19: A Patient-Reported Electronic Daily Diary in Clinical and Real-World Studies

**DOI:** 10.1101/2021.07.06.21259654

**Authors:** Diana Rofail, Nadine McGale, Joseph Im, Alissa Rams, Krystian Przydzial, Vera Mastey, Sumathi Sivapalasingam, Anna J. Podolanczuk

## Abstract

**Importance:** At the onset of the COVID-19 pandemic, there was limited understanding of symptom experience and disease progression.

**Objective:** We developed and validated a fit-for-purpose, disease-specific instrument to assess symptoms in patients with COVID-19 to inform endpoints in an interventional trial for non-hospitalized patients.

**Design:** The initial drafting of the 23-item Symptoms Evolution of COVID-19 (SE-C19) Instrument was developed based on the Centers for Disease Control and Prevention symptom list and available published literature specific to patients with COVID-19 as of Spring 2020. The measurement principles in the Food and Drug Administration patient-reported outcomes guidance and the four methodological Patient-Focused Drug Development Guidances were also considered.

**Setting:** Interviews were conducted virtually with patients recruited through a healthcare research firm.

**Participants:** Semi-structured qualitative interviews were conducted with a purposive sample of 30 non-hospitalized patients with COVID-19

**Intervention:** Interviews involved two stages: (1) concept elicitation, to obtain information about the symptoms experienced as a result of COVID-19 in patients’ own words, and (2) cognitive debriefing, for patients to describe their understanding of the SE-C19 instructions, specific symptoms, response options, and recall period to ensure the content of the SE-C19 is relevant and comprehensive. Five clinicians treating COVID-19 outpatients were interviewed to obtain their insights on symptoms experienced by patients and provide input on the SE-C19.

**Main Outcome and Measure:** Patients reported no issues regarding the relevance or appropriateness of the SE-C19 instructions, including the recall period. The comprehensiveness of the SE-C19 was confirmed against the conceptual model developed in the qualitative research. Minor conceptual gaps were revealed to capture nuances in the experience of nasal and gustatory symptoms, and systemic manifestations of sickness.

Almost all items were endorsed by patients as being appropriate and well understood. The clinicians largely approved all items, response options, and recall period.

**Conclusions and Relevance:** The qualitative research provided supportive evidence of the content validity of the SE-C19 instrument to assess the symptoms of outpatients with COVID-19 and its use in clinical trials to evaluate the benefit of treatment. Minor changes may be considered to improve conceptual clarity and ease of responding.

**Trial Registration:** R10933-10987-COV-2067 (https://clinicaltrials.gov/ct2/show/NCT04425629)

## Background

Coronavirus disease 2019 (COVID-19) is a disease caused by severe acute respiratory syndrome coronavirus 2 (SARS-CoV-2) and was declared a pandemic by the World Health Organization (WHO) in March 2020.^1^ Initial reports of common COVID-19 symptoms included fever, cough, sputum, and fatigue.^3^ By May 2020, the Centers for Disease Control (CDC) proposed additional symptoms indicative of possible COVID-19 infection: shortness of breath or difficulty breathing, muscle or body aches, headache, loss of taste or smell, sore throat, congestion or runny nose, nausea or vomiting, and diarrhea. These guidelines are consistent with other national guidelines, such as the National Health Service (NHS),^4^ and recent studies have confirmed that these symptoms can be key predictors of a positive COVID-19 test.^5^

To date, studies and guidelines have focused predominantly on hospitalizations and deaths; however, it is equally important to understand the emotional and physical effects of COVID-19 on outpatients’ lives.^6^ Currently, there are no existing patient-reported outcome measures (PROs) specific to COVID-19 symptoms. Other PROs specific to influenza and influenza-like illnesses (eg, Flu-PRO^7^) have been used in COVID-19,^8,9^ but have not been developed for or validated in this population. A targeted PRO is needed to support accurate measurement of COVID-19 symptoms, document their severity, and assess resolution.^10^ This would be particularly useful in clinical trials^11^ and real-world studies/registries in outpatients.^12^

To address this unmet need, we developed and validated a new PRO to be completed as an electronic diary: The Symptoms Evolution of COVID-19 (SE-C19). This paper summarizes the work that was conducted to (1) develop the SE-C19 and (2) evaluate its content validity in line with US Food and Drug Administration (FDA) guidance and good research practices.^13^ This paper is part of a larger body of work addressing the humanistic impact of outpatients with COVID-19, focusing on both symptoms and impacts on their daily lives (submitted for publication).

## Methods

### Stage 1: Development of the SE-C19 Questions

In early 2020, there was a limited understanding of the symptoms experienced by outpatients with COVID-19. The SE-C19 questions were developed based on the CDC symptom list^14^ and published literature available at the time.^15^ The good measurement principles outlined in the FDA PRO Guidance^13^ and the Patient-Focused Drug Development guidance^16^ formed the basis of the development.

Symptoms identified in the literature and on the CDC website were used to develop 23 questions concerning the following: feeling feverish, chills, sore throat, cough, shortness of breath or difficulty breathing, nausea, vomiting, diarrhea, headache, red or watery eyes, body and muscle aches, loss of taste or smell, fatigue, loss of appetite, confusion, dizziness, pressure or tight chest, chest pain, stomachache, rash, sneezing, sputum/phlegm, and runny nose. Patients were asked to select which of the symptoms they had experienced in the past 24 hours in an electronic diary, then rate the severity of each selected symptom at its worst in that 24-hour period on a severity scale (mild, moderate, severe). To aid interpretation of the SE-C19, additional questions were developed to assess patients’ global impressions of overall symptom severity and return to usual health/daily activities, in line with FDA recommendations.^10^ The electronic app could be accessed using a smartphone, tablet, or computer.

### Stage 2: Content Validation of the SE-C19

#### Participants

We purposively recruited 30 adult (≥18 years of age) outpatients through a healthcare research firm. All patients were recruited in the US and had a confirmed diagnosis of COVID-19, through a positive polymerase chain reaction (PCR) test, up to 21 days before the interview. Patients self-reported fever (≥100° F or feeling feverish or chills), cough, shortness of breath/difficulty breathing, change or loss of taste or smell, vomiting or diarrhea, or body/muscle aches. Patients who had been hospitalized within the past 30 days, were not fluent in English, or who were residing in nursing homes were excluded from the sample.

Interviews were conducted between September to October 2020. Before the final interviews, a target of 30 patients were recruited to achieve conceptual saturation – the point at which no new information emerges from additional interviews.^17,18^ Five independent clinicians who regularly treated patients with COVID-19 (≥5 per week) were also interviewed. These clinicians were recruited through the same healthcare research firm and Regeneron Pharmaceuticals Inc.

#### Interview Conduct

Both patient and clinician interviews (∼60 minutes) were conducted virtually by experienced qualitative researchers who received project-specific training. The semi-structured patient interview guide was divided into (1) concept elicitation and (2) cognitive debriefing.

The concept elicitation comprised open-ended questions about patients’ COVID-19 symptoms (“Can you tell me about the symptoms you have experienced with COVID-19?”). Interviewers also used structured prompts for all common symptoms of COVID-19^14^ not spontaneously mentioned. The focus of this exercise was to spontaneously elicit proximal and distal concepts associated with the COVID-19 experience, prior to cognitive debriefing of the SE-C19. This was partly in aim to assess conceptual coverage of the SE-C19, see (submitted for publication) for full account of methods and results.

During cognitive debriefing, patients used a “think aloud” method to respond to each of the questions in the SE-C19. Patients were asked to read each question aloud and verbalize their thought process while responding. When necessary, interviewers asked open-ended questions to elicit additional information about the patients’ experience responding to each question (“Tell me what you are thinking about…” or “What are you considering when you respond to this question?”).^19-21^ Once patients were given an opportunity to spontaneously provide feedback on the SE-C19, structured probes were used to elicit patients’ understanding of the instructions, clarity, and appropriateness of the instrument. Each question was reviewed to understand its relevance, ease of selecting a response, and whether the 24-hour recall period was appropriate. The objective was to ensure the intended meaning of each question of the SE-C19 was consistent with the patient’s interpretation. Patients were also asked about missing symptoms or if any questions should be removed.

Clinicians were presented with a paper copy of the SE-C19 and were asked to provide feedback on its questions (specifically any overlapping symptoms or groups of symptoms), as well as the relevance and appropriateness of the instructions, response options, and recall period.

#### Ethics

Study documents, including the protocol, demographic and health information form, interview guide, screener, informed consent form, and SE-C19, received ethical approval from the New England Independent Review Board (study number: 1291666) prior to any contact with patients. All patients completed an electronic informed consent form before the interview.

#### Data Analysis

Interviews were audio-recorded, transcribed, and anonymized. Transcripts were coded in ATLAS.ti software^22^ using an open, inductive coding approach.^22-25^ The first transcript was independently coded by three researchers; any inconsistencies in codes were discussed and reconciled. Researchers met regularly to discuss and adjust coding guidelines when necessary. The adequacy of the sample size was assessed by saturation analysis^17,18^ performed on quintiles of subsequent transcripts.

The conceptual coverage and comprehensiveness of the SE-C19 was evaluated by mapping its questions to the symptoms elicited from the patient experience. Cognitive debriefing analysis consisted of categorizing issues spontaneously reported by patients or found by probes. Structured, multi-level coding was used to report patient feedback on each question, corresponding response, instructions for completion, issues of clarity/interpretation, and any suggested changes.

## Results

### Sample Characteristics

All 30 outpatient interviews were included in the analysis. Patient ages ranged from 18–76 years, with an average age of 45 (SD±19.4). Patients were 60% female and 87% white. The interview was conducted an average of 19 days after symptoms began and an average of 12 days after COVID-19 diagnosis. Of the outpatients experiencing symptoms at the time of the interview (83%), 68% described the overall severity of their symptoms in the past 24 hours as “mild” and 32% as “moderate”. See Table 1 for full patient demographic and health information.

**Table 1.** Overview of Patient Sample Characteristics.

|  | **n = 30** |
| --- | --- |
| Age, mean (SD) | 45.03 (19.39) |
| Gender, no. (%) |  |
| Male | 12 (40) |
| Female | 18 (60) |
| No. days between diagnosis and interview,<br>mean (SD) | 12.40 (5.29) |
| No. days since symptoms began,<br>mean (SD) | 19.17 (7.96) |
| No. days between symptoms began and<br>diagnosis, mean (SD) | 6.77 (7.96) |
| General health ratings, no. (%) |  |
| Excellent | 6 (20) |
| Very good | 8 (26.7) |
| Good | 4 (13.3) |
| Fair | 9 (30) |
| Poor | 3 (10) |
| Co-morbidities*, no. (%) | 17 (57) |

\*Comorbidities across sample include: Cardiovascular disease (eg, heart failure, coronary artery disease, cardiomyopathy, pulmonary hypertension, pulmonary fibrosis), Hypertension, Diabetes (e.g., type 1, type 2, gestational), Chronic Obstructive Pulmonary Disease (COPD), Chronic kidney disease, Cancer, Stickle cell disease, Metabolic dysfunction-associated fatty liver disease, Chronic liver disease (e.g., cirrhosis), History of stroke, Thalassemia

Five clinicians were interviewed, one in Korea – where all patients with COVID-19 are hospitalized – and four in the US. The Korean clinician was informed of the focus of this study and asked to comment specifically on their experience treating mild to moderate cases. See Table 2 for full clinician backgrounds.

**Table 2.**
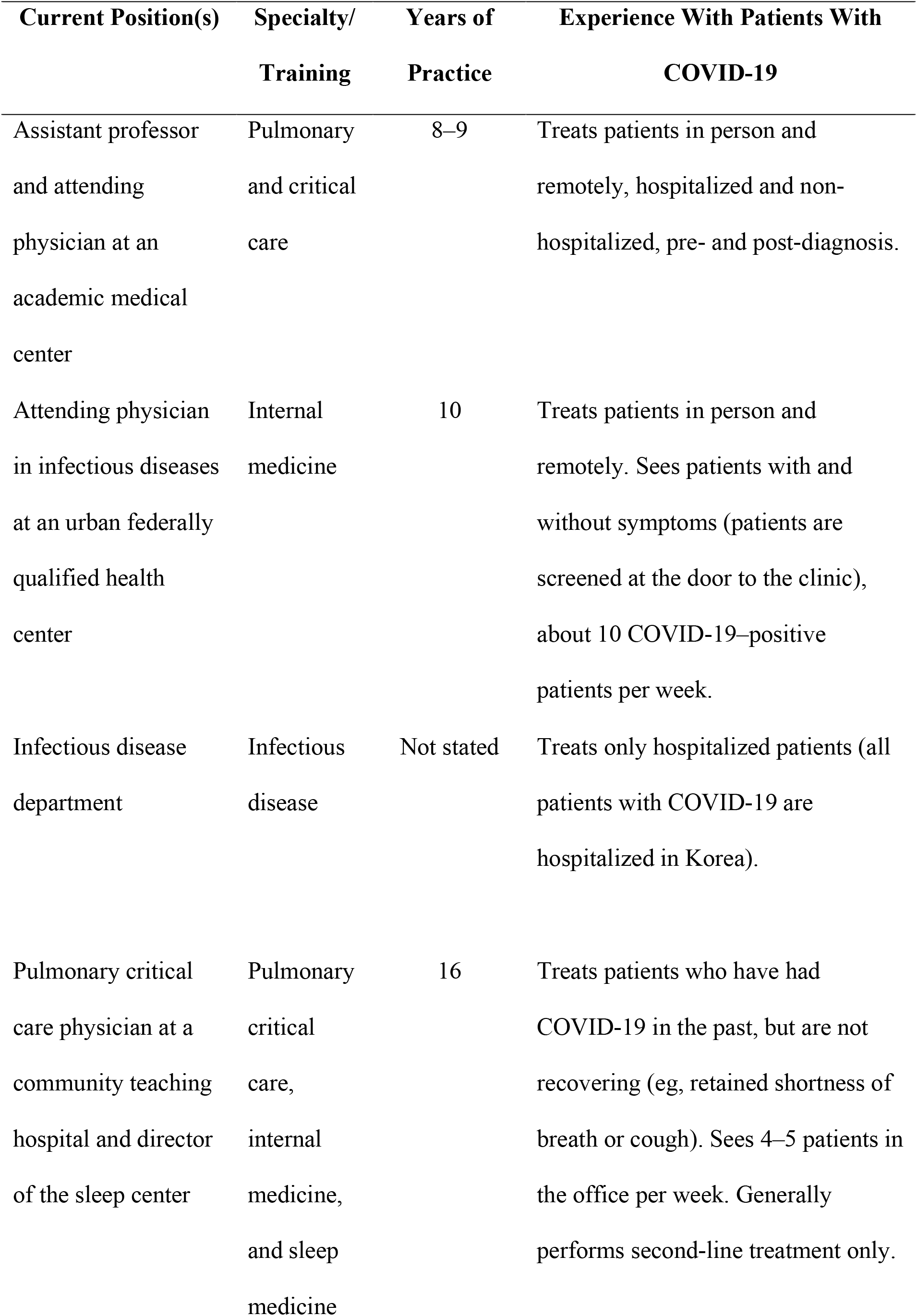
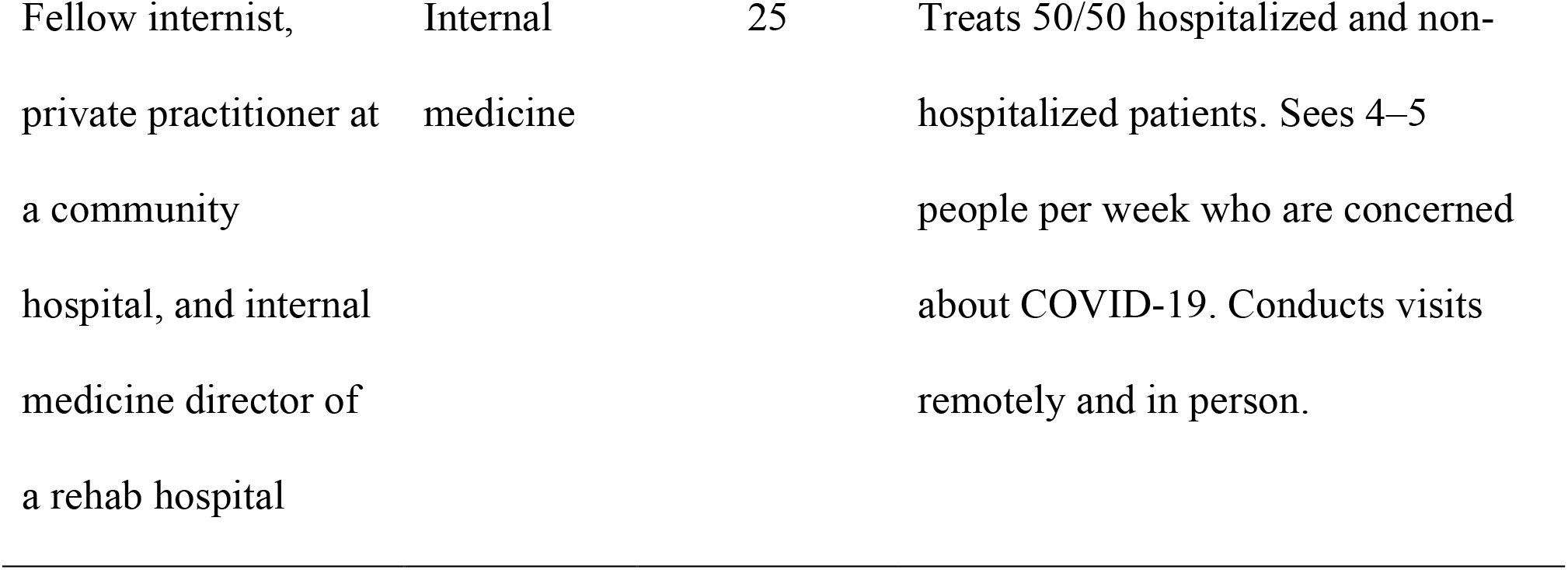
Overview of Clinician Sample Characteristics.

### Conceptual Coverage of the SE-C19 Instrument

The comprehensive list of underlying symptoms emerging from open-ended questions in the patient interviews were used for this analysis (see Table 3). The symptomatic experiences of outpatients were covered by SE-C19 questions: upper and lower respiratory tract, systemic, gastrointestinal, smell and taste, and other (skin, ocular) symptoms. Minor conceptual gaps were described by one to two outpatients regarding nuances in the experience of already captured symptoms: nasal/congestion symptoms (head congestion, stuffy nose, post-nasal drip, sinus pressure, swollen glands, earache), systemic manifestations of sickness (sweating, not feeling like their usual self, weakness, heart palpitations), and a few other rare symptoms (constipation, skin redness, numb feet).

**Table 3.**
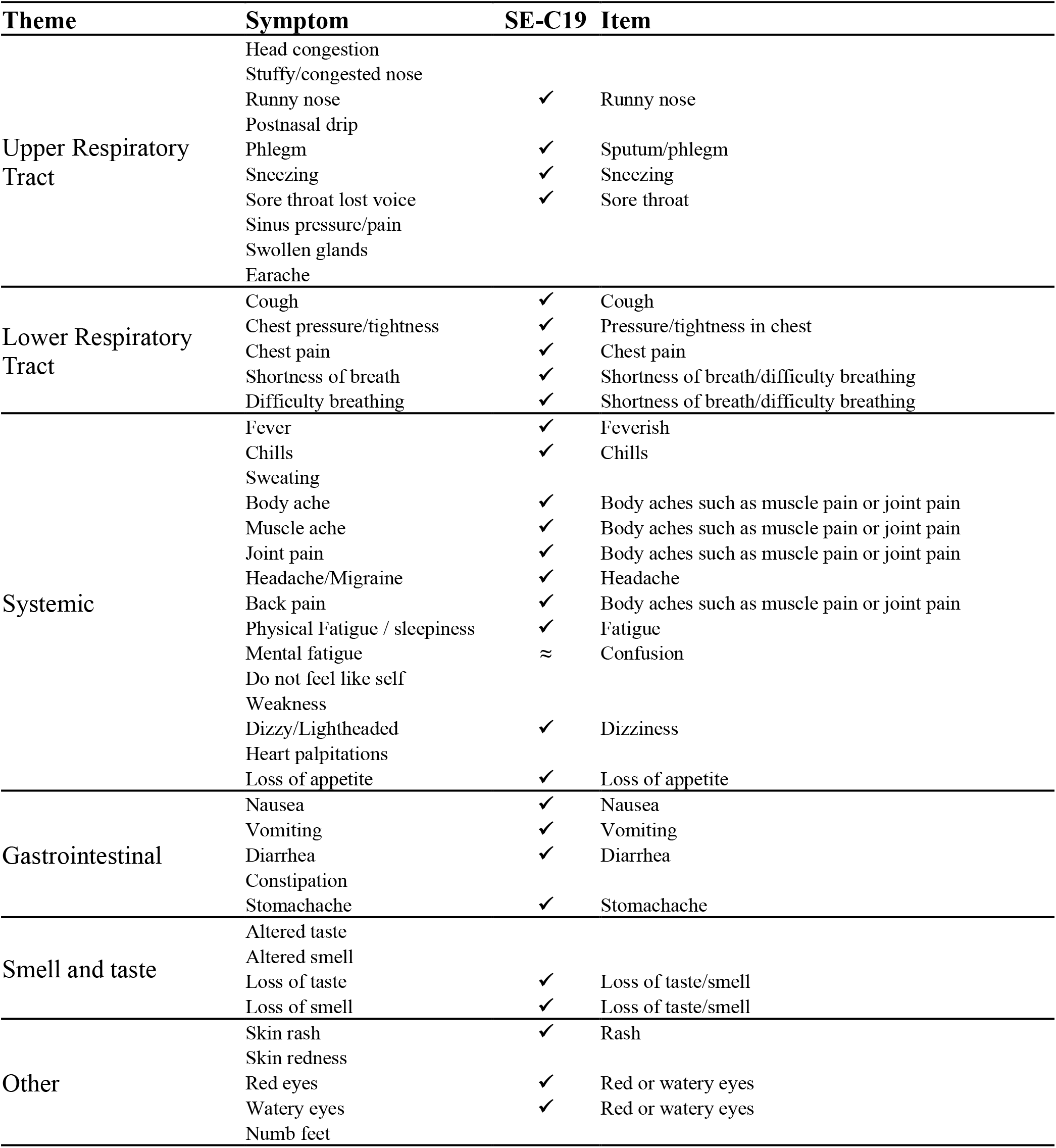
Concept-to-Item Mapping.

| Theme | Symptom | SE-C19 | Item |
| --- | --- | --- | --- |
| Upper Respiratory Tract | Head congestion |  |  |
|  | Stuffy/congested nose |  |  |
|  | Runny nose | ✓ | Runny nose |
|  | Postnasal drip |  |  |
|  | Phlegm | ✓ | Sputum/phlegm |
|  | Sneezing | ✓ | Sneezing |
|  | Sore throat lost voice | ✓ | Sore throat |
|  | Sinus pressure/pain |  |  |
|  | Swollen glands |  |  |
|  | Earache |  |  |
| Lower Respiratory Tract | Cough | ✓ | Cough |
|  | Chest pressure/tightness | ✓ | Pressure/tightness in chest |
|  | Chest pain | ✓ | Chest pain |
|  | Shortness of breath | ✓ | Shortness of breath/difficulty breathing |
|  | Difficulty breathing | ✓ | Shortness of breath/difficulty breathing |
| Systemic | Fever | ✓ | Feverish |
|  | Chills | ✓ | Chills |
|  | Sweating |  |  |
|  | Body ache | ✓ | Body aches such as muscle pain or joint pain |
|  | Muscle ache | ✓ | Body aches such as muscle pain or joint pain |
|  | Joint pain | ✓ | Body aches such as muscle pain or joint pain |
|  | Headache/Migraine | ✓ | Headache |
|  | Back pain | ✓ | Body aches such as muscle pain or joint pain |
|  | Physical Fatigue / sleepiness | ✓ | Fatigue |
|  | Mental fatigue | ≈ | Confusion |
|  | Do not feel like self |  |  |
|  | Weakness |  |  |
|  | Dizzy/Lightheaded | ✓ | Dizziness |
|  | Heart palpitations |  |  |
|  | Loss of appetite | ✓ | Loss of appetite |
| Gastrointestinal | Nausea | ✓ | Nausea |
|  | Vomiting | ✓ | Vomiting |
|  | Diarrhea | ✓ | Diarrhea |
|  | Constipation |  |  |
|  | Stomachache | ✓ | Stomachache |
| Smell and taste | Altered taste |  |  |
|  | Altered smell |  |  |
|  | Loss of taste | ✓ | Loss of taste/smell |
|  | Loss of smell | ✓ | Loss of taste/smell |
| Other | Skin rash | ✓ | Rash |
|  | Skin redness |  |  |
|  | Red eyes | ✓ | Red or watery eyes |
|  | Watery eyes | ✓ | Red or watery eyes |
|  | Numb feet |  |  |

Most symptoms were explicitly mapped to one SE-C19 question, meaning the question (e.g., feverish) was directly associated with a patient-reported symptom (e.g., fever). One question in the SE-C19 (“confusion”) did not directly map onto one experience described by outpatients; however, outpatients did report feeling groggy, brain fog, not feeling as mentally sharp as they previously were, and mental fatigue. Additionally, the SE-C19 addresses both smell and taste in one question, but outpatients described alterations in these senses as different experiences.

### Cognitive Debriefing of the SE-C19 Instrument

The instructions of the SE-C19 were relevant and easy to understand, as were the 24-hour recall period and response options. Very few issues in understanding were reported for the questions. The most frequent issue concerned “sputum/phlegm” (n = 11); outpatients were unfamiliar with the term “sputum”, but easily understood the term “phlegm”.

> “I didn’t really know what sputum meant, but I saw phlegm, so that’s why I checked it.” (Male, 18–25)

Four outpatients felt that it was difficult to provide an accurate answer to the question “loss of smell/taste” if they were only experiencing a loss of smell or taste, but not both, or if they experienced an alteration in their senses as opposed to complete loss.

> “I thought they would go together, but I couldn’t smell first and I could still taste.” (Female, 18–25)

However, other outpatients experienced these at the time same time and thought they belonged together conceptually (n = 7).

> “I don’t suppose there’s a difference. They both are blocked, both of them at the same time, so I feel that they’re connected.” (Female, 26–30)

Two outpatients reported a response problem for the question “body aches such as muscle pain or joint pain” due to their aching experience not being specific to muscle or joint pain. One patient had difficulty with choosing between “moderate” and “severe” for the question “shortness of breath/difficulty breathing”, as they indicated that, though they had experienced some instances that could be considered “severe”, their symptoms were generally moderate.

Differences between “runny nose” (included in the SE-C19) and “stuffy nose” (not included in SE-C19), were probed. Five outpatients believed these were different, but four other outpatients saw them as connected experiences.

> “Usually stuffy nose starts from runny nose. So, they are connected together.” (Female, 60–65)

“Nausea” and “vomiting” were confirmed as conceptually different by all outpatients who endorsed these symptoms (n = 22). Outpatients also believed that “pressure/tightness in chest” and “chest pain” were conceptually different experiences and agreed that they should remain as separate questions (n = 18).

> “I personally feel like they’re different because I do have a lot of chest pressure, but, really, the only time I experienced pain in my chest was just from the constant coughing.” (Female, 26–30)

Almost all outpatients (n = 28) thought the symptom list was comprehensive in reflection of their own experience, and the only symptoms listed as possible additions were “earache” and “voice loss”, each only reported by one outpatient.

### Clinician Interviews

The clinicians supported the importance and relevance of all symptoms that were currently combined into a singular question. However, one indicated that “body aches such as muscle pain and joint pain” could be separated into two questions if the scale were to be administered long-term, as joint pain can linger more than other body pains. Furthermore, clinicians felt that the term “body aches” did not add any specificity to the question and recommended removing it. Regarding “sputum/phlegm”, clinicians agreed that sputum and phlegm are not clinically different but stated that patients are not often familiar with the term “sputum”. Finally, “shortness of breath/difficulty breathing” was not considered clinically different in mild cases, but clinicians felt that “shortness of breath” was a more relevant, general term than “difficulty breathing”.

From the clinicians’ perspectives, some questions in the SE-C19 were considered less common in outpatients with COVID-19: runny nose, sneezing, confusion, dizziness, stomachache, rash, and red/watery eyes. Clinicians reported that “confusion” was typically seen in hospitalized or older patients with severe cases of COVID-19. Minor conceptual gaps that were identified in the concept-to-item analysis based on outpatient results were not identified by clinicians as core to the outpatient experience of COVID-19.

The clinicians endorsed the response options regarding the severity level of each question (mild, moderate, severe). Four of the five clinicians approved a 24-hour recall period for completing the SE-C19; the fifth clinician did not provide specific feedback on the recall period. One clinician felt that if the diary is successfully administered daily, it will likely be able to capture change in patients’ symptomatic experiences. If administered less often, the symptom experience of COVID-19 would not be captured adequately.

## Discussion

The purpose of this study was to develop the SE-C19, a new PRO instrument that is appropriate, easy to understand, and comprehensive for measuring symptom evolution in outpatients with COVID-19. To our knowledge, there are no fit-for-purpose PROs available. Although the Flu-PRO has been used as a PRO to measure COVID-19,^8,9^ it has not been validated in outpatients with the disease. The development of the SE-C19 addresses the important unmet need for a standardized method for evaluating COVID-19 symptoms.

The questions of the SE-C19 were developed based on the symptoms reported in clinical literature and listed on the CDC website at the time (May 2020), which provides face validity in tracking symptom onset and recovery. The current paper provides further empirical evidence to support clinical guidelines, based on data from both outpatients and clinicians. In addition to the face validity, this research was important to establish the content validity of the SE-C19 in two ways: first, to confirm the coverage of the symptomatic experience of outpatients based on a comprehensive list of concepts elicited before outpatients saw the SE-C19; second, to confirm the questions were appropriate and understood through the cognitive debriefing.

The SE-C19 covered the symptoms that outpatients described as part of their experience. Minor conceptual gaps described by one to two outpatients revealed nuances in the experience of nasal/congestion or systemic manifestations of sickness. Few additional symptoms (constipation, skin redness, numb feet) were reported by individual outpatients (Table 3). Once outpatients reviewed all questions in the SE-C19, they agreed that no important symptoms were missing.

The symptoms proposed in the FDA guidance for COVID-19 clinical trials released after the development of the SE-C19,^16^ as well as the symptoms listed by the CDC,^14^ WHO,^1^ and NHS,^4^ are all measured by the SE-C19. The 24-hour recall period and use of a 4-point scale are also supported by the FDA guidance.

The cognitive debriefing of the SE-C19 confirmed that most questions were appropriate, well understood, and easy to answer. The two major findings of the cognitive debriefing were: (1) the word “sputum” in the “sputum/phlegm” question was not understood by some outpatients, but this did not impact their ability to respond, as “phlegm” was understood; (2) some outpatients reported difficulty answering the question about “loss of taste/smell” since they considered these different symptoms. Some outpatients also reported that “altered” taste was different from “lost” taste.

Future studies could consider minor adjustments to the SE-C19 to improve the conceptual coverage of nuances in the symptomatic experience and improve the understanding of the following questions: adding one question related to congestion/stuffy nose, separating “loss of taste/smell” into two questions that further evaluate altered and lost concepts separately, rewording “confusion” to “brain fog” to more accurately reflect the outpatient experience, rewording “body aches such as muscle pain and joint pain” to “body or muscle aches”, and removing “sputum” from “sputum/phlegm”.

The current study has a few limitations. Some outpatients were no longer experiencing symptoms at the time of the interviews, as they were completed an average of 12 days after their positive diagnosis. However, outpatients had vivid memories of their past symptoms and were able to provide detailed feedback for the SE-C19. Due to the small sample size of the current study, further research should be conducted to confirm the validity and reliability of all the SE-C19 questions, using classical and modern test theory to support the scoring of the measure and define responders. This will also provide evidence for the minor adjustments suggested based on the current sample’s feedback. Upon completion of further analyses, this provides a valid, evidence-based PRO assessment of COVID-19 that can be considered for future use by patients and healthcare professionals. While the current study focused on outpatients in the acute phase of COVID-19, additional research could further explore the symptom trajectory and impact on daily lives in long-term COVID.^26^ In this context, the SE-C19 could be a useful tool to better understand persistent symptoms.

## Conclusion

The current study provides supportive evidence for the content validity of the SE-C19 to assess the symptoms of outpatients with COVID-19 and its use in clinical trials to evaluate treatment benefit, with suggestions for minor adjustments. The SE-C19 addresses the need for a comprehensive PRO assessing the symptoms of outpatients with COVID-19, their severity, and degrees of recovery, especially with the growing number of genetic variants of SARS-CoV-2.^27^ It can also be used to assess patient burden in those experiencing “long COVID”.^26^

To conclude that the amended version of the SE-C19 is fit for purpose in outpatient clinical research, the next phase will involve quantitative evaluation of its psychometric performance, including a scoring algorithm, a strategy to derive endpoints from daily data, and the determination of within-patient meaningful change.

## Supporting information

AP ICMJE

AR ICMJE

DR ICMJE

JI ICMJE

KP ICMJE

NM ICMJE

SS ICMJE

VM ICMJE

## Data Availability

Not applicable.

## Acknowledgments

We thank the study participants, their families, and the clinicians involved in this trial. The authors also thank Prime, Knutsford, United Kingdom for manuscript formatting and copy-editing suggestions.

## Author Contributions

DR contributed to conceptualization (including original and refined versions of the SE-C19, as well as initial conception and design of the study), funding acquisition, methodology, project administration, supervision, visualization, and writing (original draft, review and editing).

NM contributed to conceptualization, data curation, methodology, formal analysis, project administration, supervision, visualization, and writing (review and editing).

JI contributed to conceptualization, supervision, and writing (review and editing).

AR contributed to data curation, methodology, formal analysis, visualization, and writing (original draft, review, and editing).

KP contributed to data curation, methodology, formal analysis, visualization, and writing (original draft, review, and editing).

VM contributed to conceptualization, supervision, and writing (review and editing). SS contributed to conceptualization, supervision, and writing (review and editing). AP contributed to conceptualization, supervision, and writing (review and editing).

## Conflict of Interest Disclosures

DR is a Regeneron Pharmaceuticals, Inc. employee/stockholder and former Roche employee, and current stockholder. JI and VM are Regeneron Pharmaceuticals, Inc. employees/stockholders. NM, AR, and KP are employees of Modus Outcomes and consulted for Regeneron Pharmaceuticals, Inc. SS is an Excision BioTherapeutics employee/stockholder and former Regeneron Pharmaceuticals, Inc. employee, and current stockholder. AP has received consulting fees from Regeneron Pharmaceuticals, Inc., honoraria from NACE (CME), and has participated in an advisory board for Boehringer Ingelheim.

## Funding/Support

This research was sponsored and paid for by Regeneron Pharmaceuticals, Inc.

## Role of the Funding Source

This study was funded by Regeneron Pharmaceuticals, Inc.

## Data Sharing

Not applicable.

## Notes

### Competing Interest Statement

DR is a Regeneron Pharmaceuticals, Inc. employee/stockholder and former Roche employee, and current stockholder. JI, SS, and VM are Regeneron Pharmaceuticals, Inc. employees/stockholders. AP has received consulting fees from Regeneron Pharmaceuticals, Inc., honoraria from NACE (CME), and has participated in an advisory board for Boehringer Ingelheim. NM, AR, and KP are employees of Modus Outcomes. NM, AR, and KP consulted for Regeneron Pharmaceuticals, Inc.

### Clinical Trial

NCT04425629

